# Machine Learning-Based Meningioma Location Classification Using Vision Transformers and Transfer Learning

**DOI:** 10.1101/2025.09.22.25336380

**Authors:** Jathin K. Bande, Ethan Thomas Johnson, Rajesh Banderudrappagari

## Abstract

**Purpose:** In this study, we aimed to use advanced machine learning (ML) techniques, specifically transfer learning and Vision Transformers (ViTs), to accurately classify meningioma in brain MRI scans. ViTs process images similarly to how humans visually perceive details and are useful for analyzing complex medical images. Transfer learning is a technique that uses models previously trained on large datasets and adapts them to specific use cases. Using transfer learning, this study aimed to enhance the diagnostic accuracy of meningioma location and demonstrate the capabilities the new technology.

**Approach:** We used a Google ViT model pre-trained on ImageNet-21k (a dataset with 14 million images and 21,843 classes) and fine-tuned on ImageNet 2012 (a dataset with 1 million images and 1,000 classes). Using this model, which was pre-trained and fine-tuned on large datasets of images, allowed us to leverage the predictive capabilities of the model trained on those large datasets without needing to train an entirely new model specific to only meningioma MRI scans. Transfer learning was used to adapt the pre-trained ViT to our specific use case, being meningioma location classification, using a dataset of 1,094 images of T1, contrast-enhanced, and T2-weighted MRI scans of meningiomas sorted according to location in the brain, with 11 different classes.

**Results:** The final model trained and adapted on the meningioma MRI dataset achieved an average validation accuracy of 98.17% and a test accuracy of 89.95%.

**Conclusions:** This study demonstrates the potential of ViTs in meningioma location classification, leveraging their ability to analyze spatial relationships in medical images. While transfer learning enabled effective adaptation with limited data, class imbalance affected classification performance. Future work should focus on expanding datasets and incorporating ensemble learning to improve diagnostic reliability.

## 1 Introduction

Meningiomas are tumors that arise from the meninges, and they account for approximately 30% of all brain tumors. Although often non-cancerous (benign), they can cause life-threatening effects on the brain due to the different locations in which the tumor may arise, some of which are heavily affected by the tumor’s presence (Ostrom et al., 2019). Meningioma tumors must be diagnosed early, and the tumor location also needs to be precisely identified. This is a necessity for further surgical intervention.

Recent advancements in artificial intelligence (AI) have demonstrated the potential of machine learning (ML) to bolster diagnostic accuracy in medical imaging. This pertains especially in the fields of radiology and oncology (Esteva et al., 2017). This is due to ML’s capability of making patient diagnoses less variable and easier to derive. Convolutional Neural Networks (CNNs) have been invaluable to AI in medicine and have proved to be immensely helpful in achieving accurate computer-led diagnosis. However, evidence has shown that CNNs and other such ML models may not effectively look at complex medical images, meaning that important global and contextual information is commonly missed. (Litjens et al., 2017).

To solve this issue, vision transformers (ViTs) may be used, as they can identify global relationships within medical imaging. Due to this, ViTs may rival CNNs in medical image diagnosis (Dosovitskiy et al., 2020). ViTs utilize self-attention mechanisms, which were initially developed for natural language processing tasks, to analyze images in a non-sequential manner. Essentially, ViTs split up an image into many smaller parts while still relating the smaller parts to the entire, global image. This allows them to capture complex spatial dependencies across an entire image. As a result, ViTs are highly effective at analyzing complex images, which in this case includes brain MRI scans. Initial studies have shown promise in areas such as tumor detection and segmentation using the relatively new technology (Azad et al., 2023).

Transfer learning is a technique that adapts a previously trained model to a new but related task. This technique has been explored in medical imaging tasks, specifically when there is not an abundance of training data available. (Yosinski et al., 2014). By starting with a model trained on a large dataset of general images, transfer learning allows researchers to build accurate models with less data and less training time. Transfer learning may not only accelerate model training, but could also improve performance in specialized domains, such as the classification and localization of tumors in MRI scans (Raghu et al., 2019).

In this study, a key objective was to explore the application of Vision Transformers and transfer learning for meningioma location classification in MRI images. A pre-trained Vision Transformer model was fine-tuned on a dataset of meningioma MRI scans to assess its performance in classifying tumor locations. This research aims to contribute to the growing field of machine learning in medical imaging. It aims to demonstrate the potential of Vision Transformers and transfer learning in developing accurate, reliable diagnostic tools.

## 2 Materials & Methods

### 2.1 Data

All data used in the paper is available in the public domain.

#### 2.1.1 Meningioma Dataset

A dataset consisting of 1,094 meningioma MRI scans was used to train and evaluate the machine learning model adapted from the pre-trained ViT. The dataset had 11 different classes of varying tumor locations within the brain, and was a collection of T1, contrast-enhanced, and T2-weighted MRI images. These MRI images were without any type of marking or patient identification, and they were interpreted by radiologists and provided for study purposes. Images were separated into classes including anterior fossa, cavernous, frontal, infratentorial, intracisternal, parafalcine, parietal, petroclival, suprasellar, supratentorial, and temporal locations (Feltrin, 2023). Multiple image augmentations were applied to the images to generate more robust training data.

**Figure 1.**
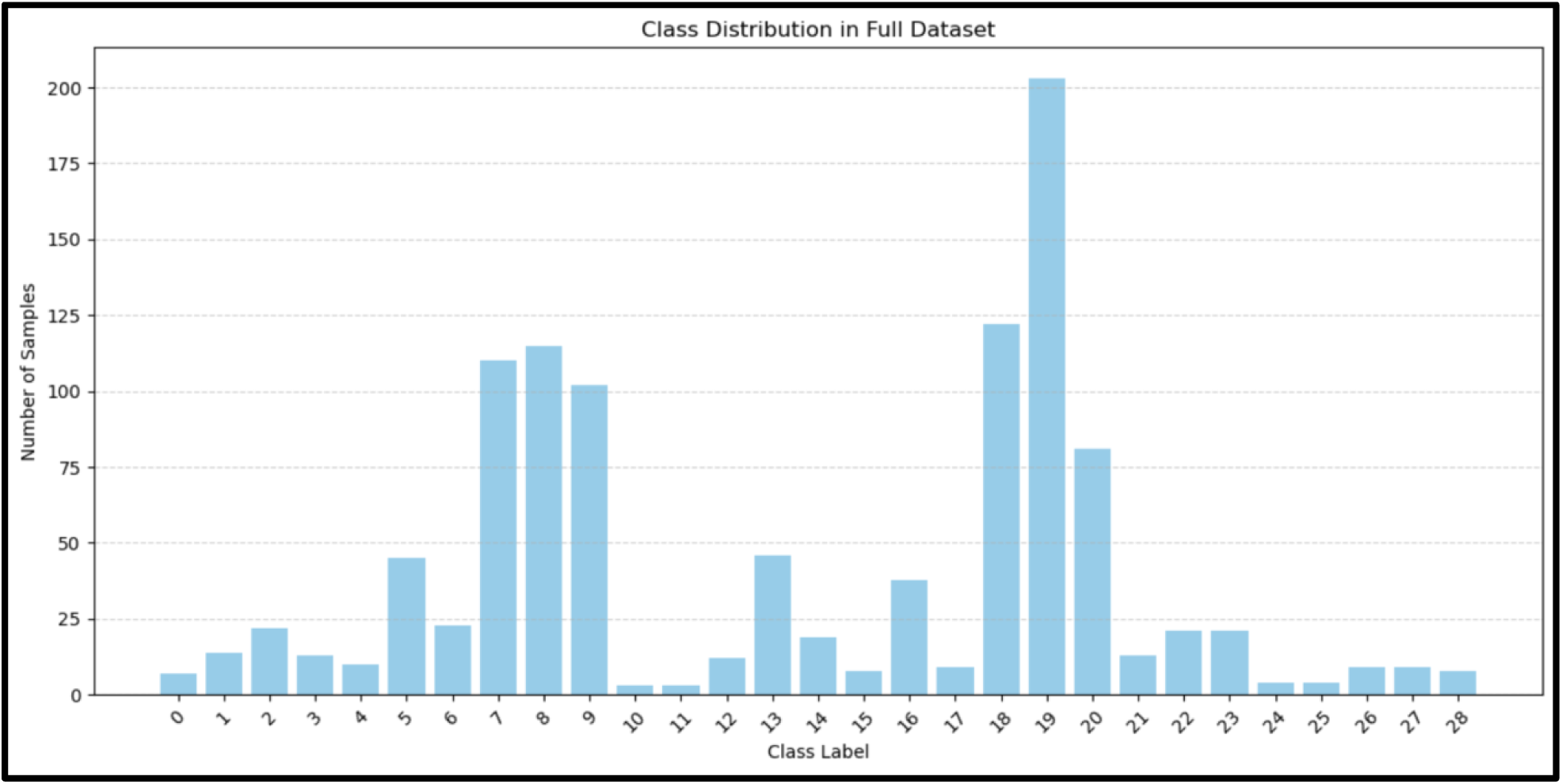
A histogram representing the class distribution of the full dataset prior to data modification, consisting of 29 separate classes of meningioma MRI scans based on specific locations.

**Figure 2.**
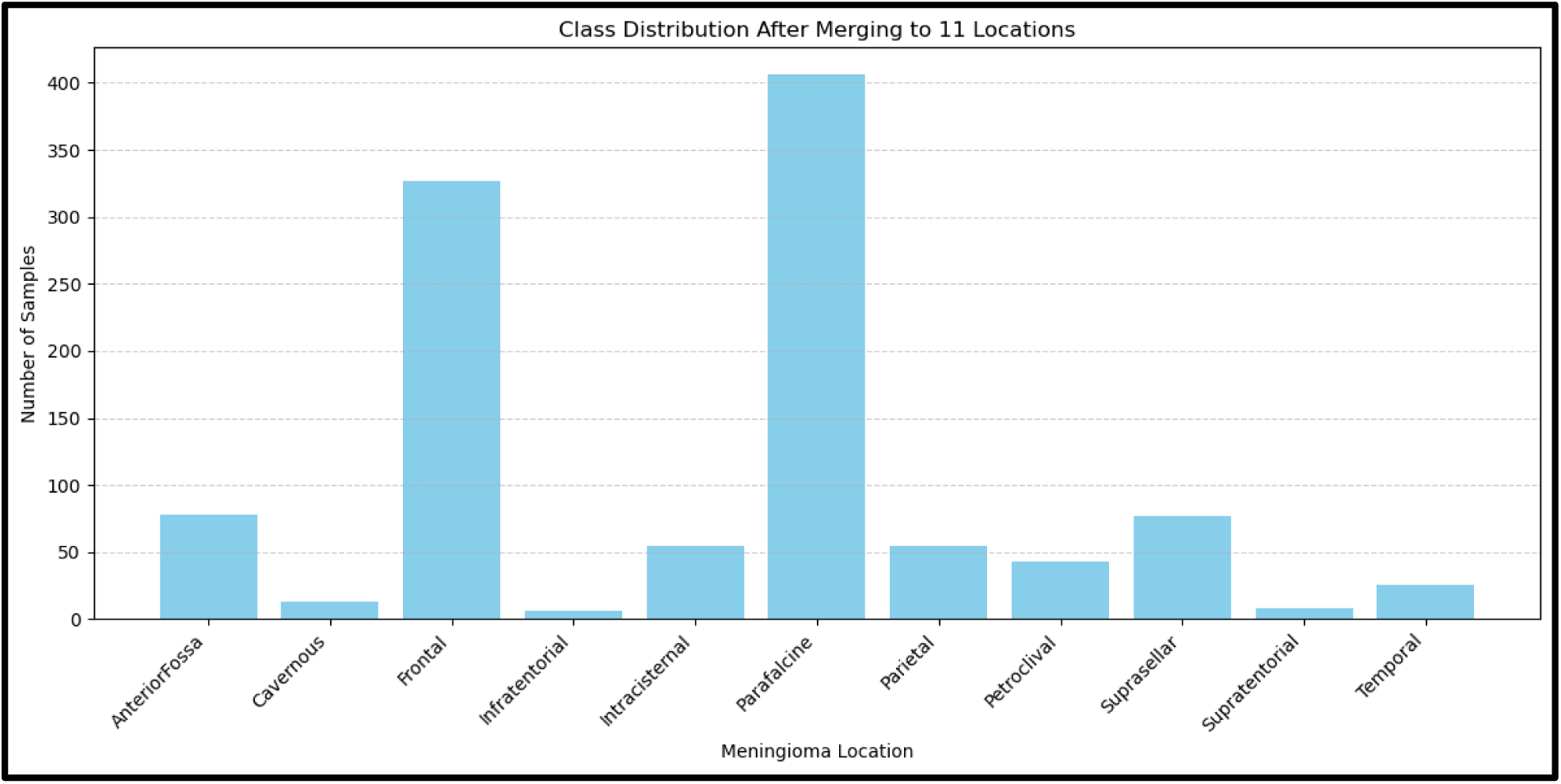
A histogram representing the class distribution of the meningioma MRI scans after merging to 11 classes based on general tumor locations.

**Figure 3.**
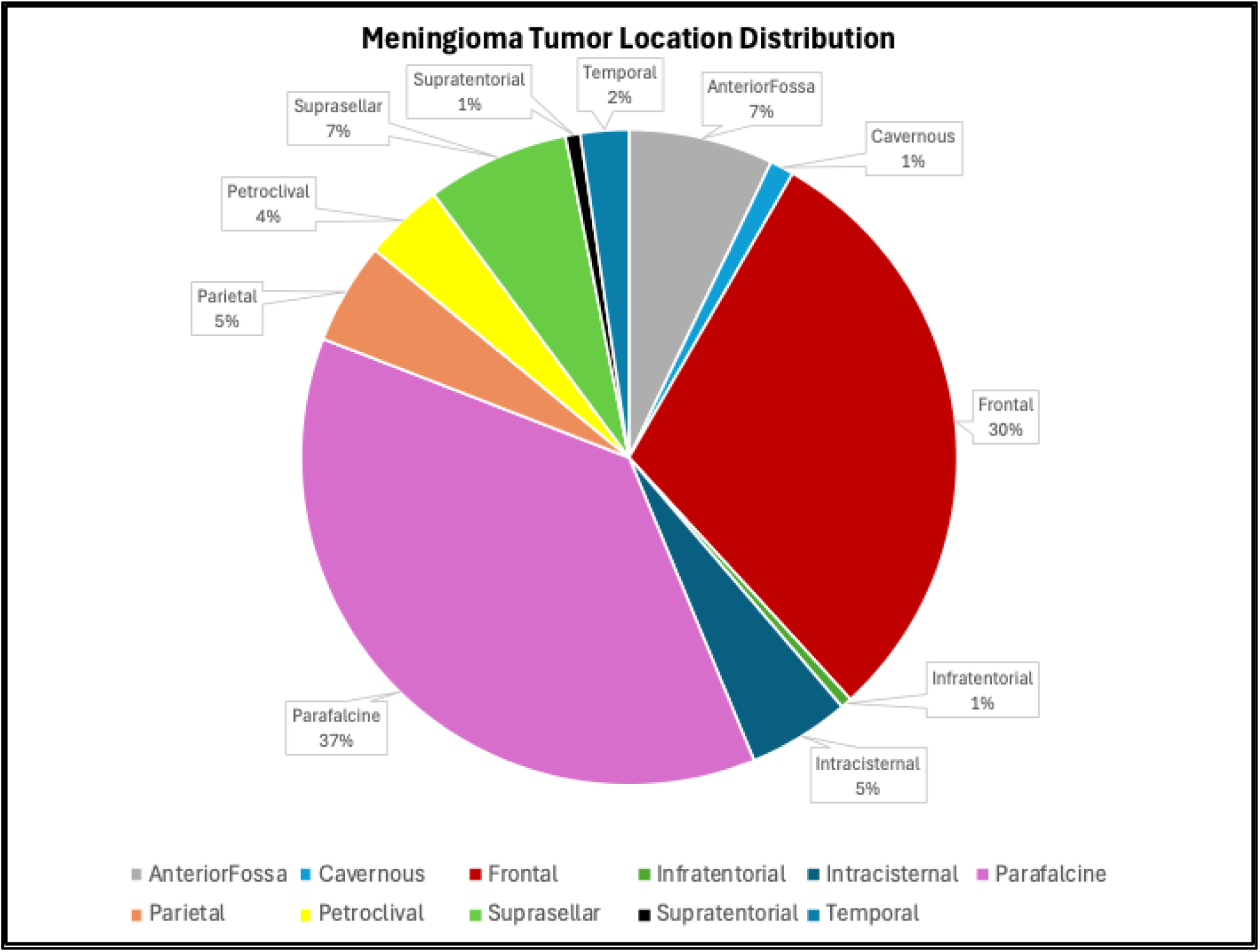
A pie chart representing the percentage distributions of the meningioma MRI scans after merging to 11 classes based on general tumor locations.

### 2.2 Model Creation

#### 2.2.1 Original Vision Transformer

The ViT model (vit-base-patch16-224) was pre-trained on ImageNet-21k (14 million images, 21,843 classes) and further fine-tuned on ImageNet 2012 (1 million images, 1,000 classes) (Dosovitskiy et al., 2020). All input images were resized to 224×224 resolution.

#### 2.2.2 Transfer Learning for Meningioma

The meningioma dataset was split into training, validation, and testing subsets, which constituted 80%, 10%, and 10% of the data, respectively. To ensure efficient model training, a dictionary was used to map numerical identification numbers to the scans. Next, all the meningioma scans were converted to grayscale, as our dataset contained both color and gray images. The images were then further processed and converted to tensor format to ensure that the data met the ViT model’s input requirements of images of 224×224 resolution, as well as 3 channels in each input image. The model was fine-tuned using 20 epochs and a learning rate of 3 × 10^−4^, with early stopping enabled with a patience of 3 epochs. 5-Fold cross validation was used for reproducibility of the results, and the best model from the 5 folds was saved. Training metrics, including accuracy and loss, were logged throughout the training process. Out of the total 85,020,957 parameters in the ViT, only 22,301 parameters in the classification layer of the ViT were trainable. This focused the computational resources while training on learning features specific to meningioma classification. The transformed model was then evaluated.

## 3 Statistical Analysis

Python libraries including NumPy, Pandas, and Scikit-Learn were used for statistical analysis. Performance metrics included evaluation accuracy, training loss, validation loss, accuracy, precision, recall, and f1-score were used for performance evaluation.

## 4 Results

The final model trained and adapted on the meningioma MRI dataset achieved an average validation accuracy of 98.17% across the 5 folds, and a test accuracy of 89.95%.

**Figure 4.**
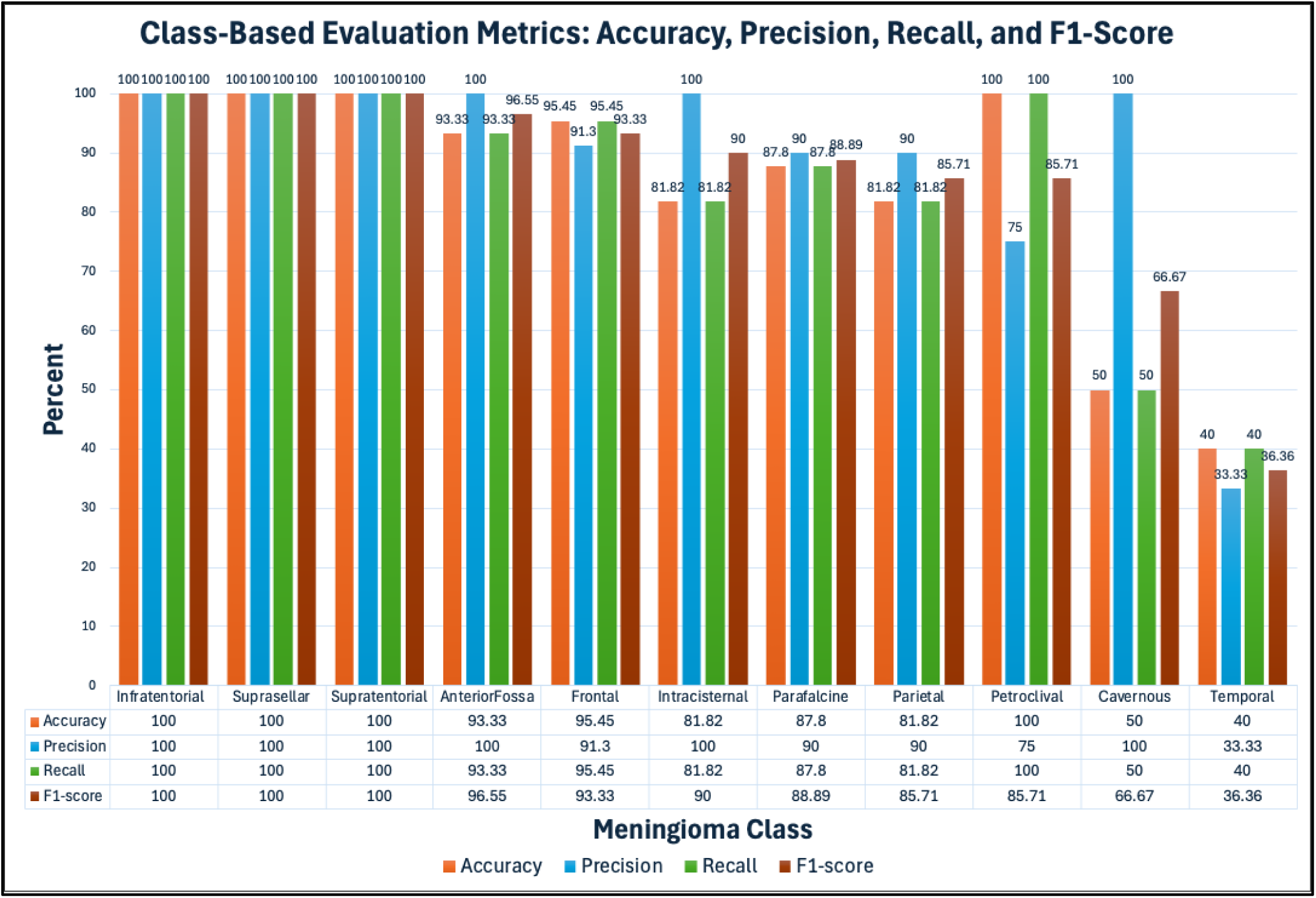
A histogram showing the class-based evaluation metrics of the model, including accuracy, precision, recall, and F1-score, along with a table showing the numerical metrics more clearly.

**Figure 5.**
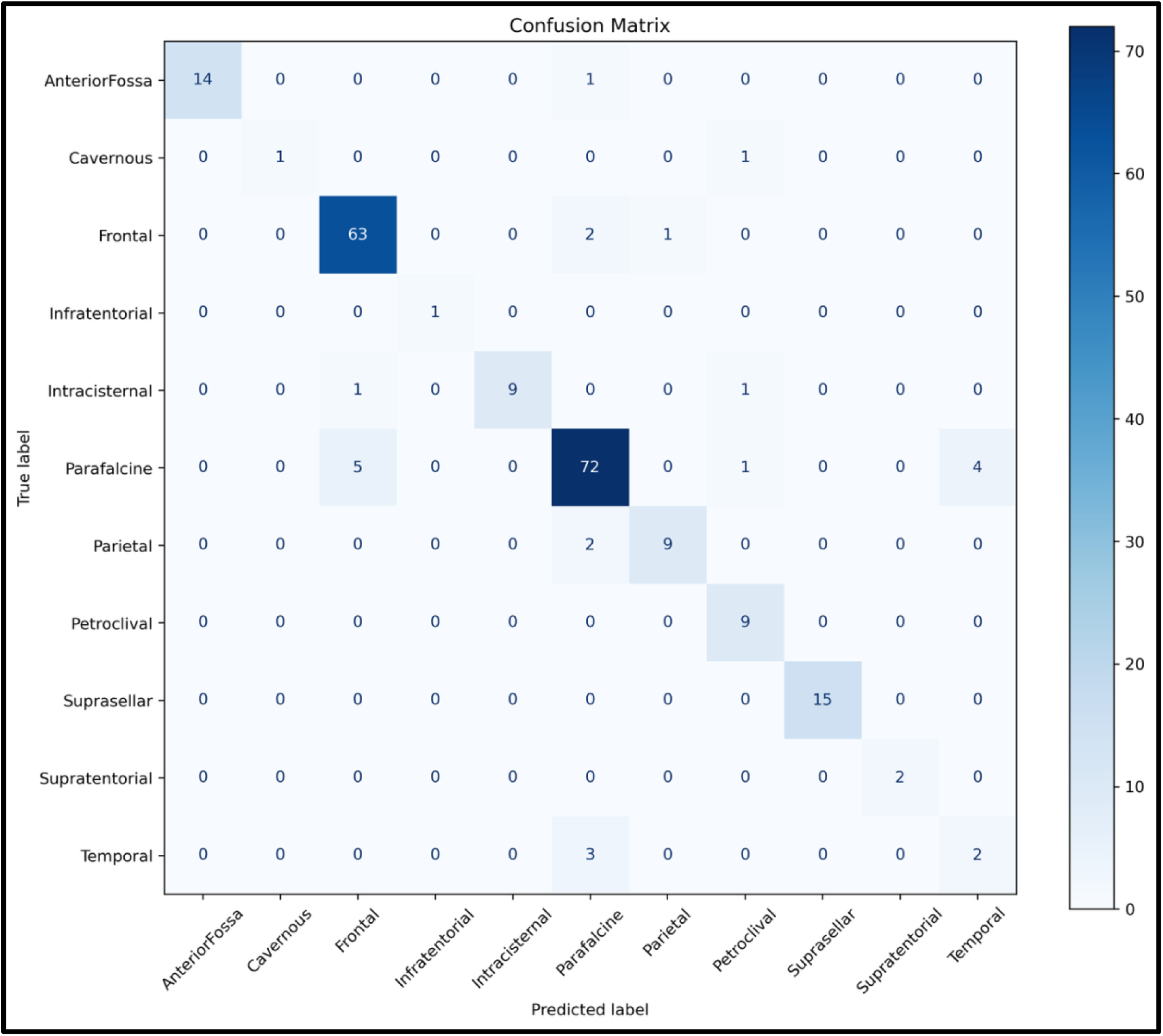
A 11×11 confusion matrix of the model’s predictions on the test set, showing the true labels vs. the model’s predicted labels.

**Figure 6.**
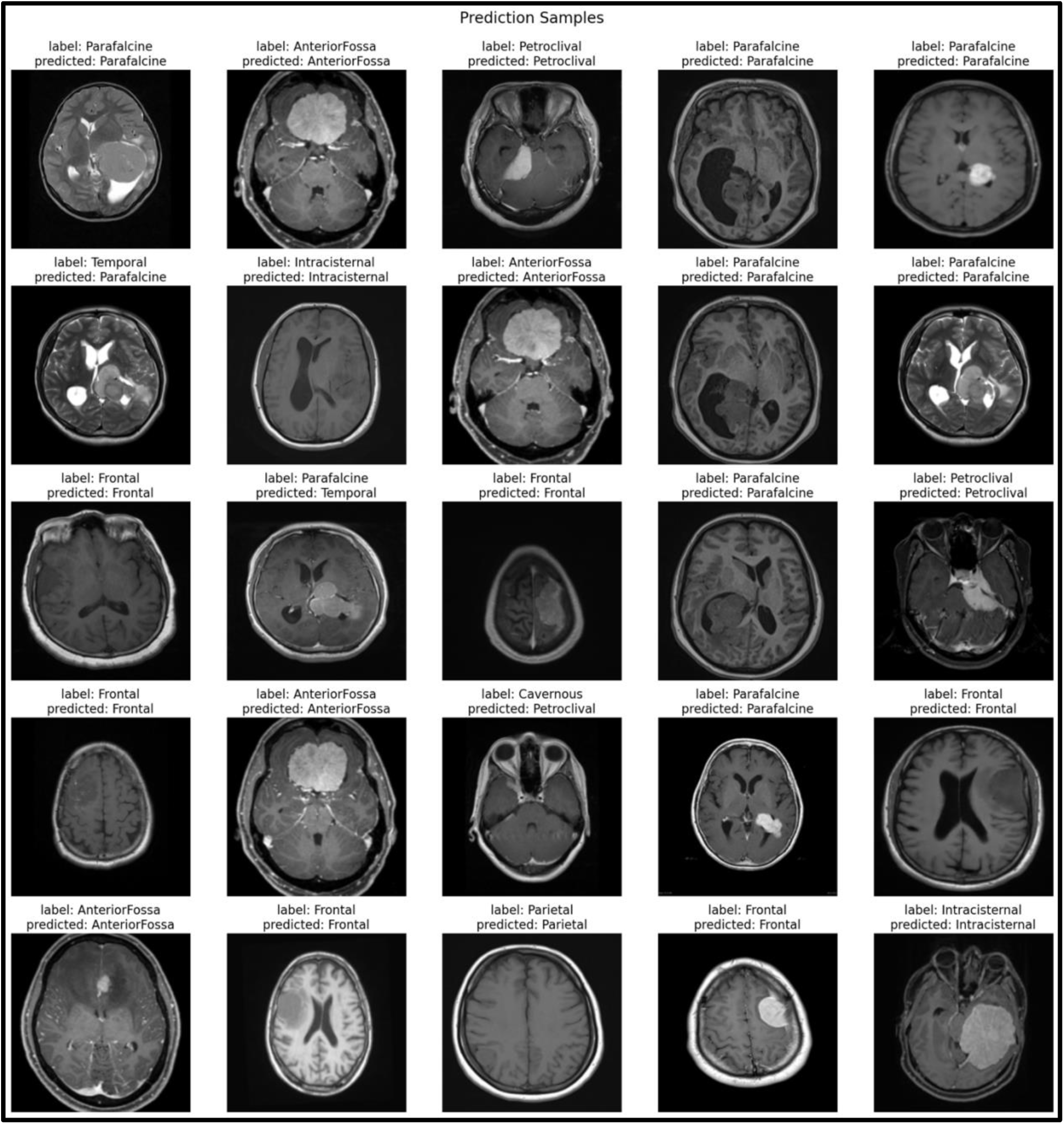
A representation of the model’s predictions for randomly selected MRI images from the testing set, showing both their true and predicted classifications.

## 5 Discussion

### 5.1 Summary and Contextualization of Findings

The State-of-the-art level average accuracy in classifying meningioma location across 11 collated classes produced by our fine-tuned Vision Transformer (ViT) highlights the potential for ViTs to be implemented in a larger diagnostic pipeline in which location classification can be utilized by clinicians to more efficiently and accurately assess meningioma prognosis and treatment plans. Prior literature in DL models for neuroimaging largely focus on brain tumor detection and classification, whereas anatomical localization has received little attention in comparison. These past implementations of DL models, primarily Convolutional Neural Networks (CNNs), have presented with comparable accuracies in detection, classification, and grading to the anatomical localization by our fine-tuned ViT. Despite most DL models in neuro-oncology using CNN architectures, as of late there have been an increase in the implementation of ViTs due to their global attention mechanisms which provide the benefit of reduced inductive bias as ViTs can better understand local tumor characteristics as well as their spatial context within the entire brain.

Our study shows how instrumental deep learning models employing attention mechanism can understand and classify tumor location. Past work has expanded upon the use of various attention mechanisms on tumor classification. Huang et al. (2025) explored the benefits of using a squeeze-and-excitation (SE) attention mechanism on fine-tuned neural network for tumor classification, specifically on meningiomas, gliomas, pituitary tumors, and no tumor. SE attention was employed in the study due to its ability to adaptively adjust feature channel weights to enhance informative channels and suppress noisy ones. SE attention works through global average pooling of each channel which yields a vector the size of the number of the filters. Then in the excitation stage this vector is fed to a small bottleneck multilayer perceptron (MLP) which produces weights ranging from 0 to 1 for each channel. Finally, the weights generated by the MLP are used to multiply the original feature maps which prioritizes important channels and repress less relevant channels. The ResNet50V2 using SE attention used in the study was shown to outperform a base model by 5.8% in accuracy achieving a classification accuracy of 98.4%. Additionally, the ResNet50V2 with SE attention showed an AUC of 0.999 as well as statistically significant increases in classification accuracy for meningioma and pituitary tumors. These results were elucidated by Huang & Prakash (2025) which finds that a SE Convolutional Block Attention Module achieved a classification accuracy of 98.4% and an AUC of 1.00 in differentiating between the same classes as the previous study. A key future direction of both studies was exploring vision transformers as a means for superior feature extraction.

Such exploration has already been established by Reddy et al. (2024) who ran a comparative analysis between various fine-tuned vision transformers (FTVTs) and a handful of well-established deep learning models like ResNet50, MobileNet-V2, and EfficientNet-B0 for classifying glioma, meningioma, pituitary, and no tumor. Among all the models, FTVT-L16, a large FTVT processing 16×16 patches, achieved the highest classification accuracy of 98.70%, outperforming ResNet-50 (96.5%), EfficientNet-B0 (95.1%), and MobileNet-V2 (94.9%). All the FVTV models had higher classification accuracies than the established model. Notably, even the lowest-performing FTVT model, FTVT-B32, a base FVTV processing 32×32 patches, surpassed the best-established model, ResNet-50, by 0.37%, with an accuracy of 96.87%.

### 5.2 Clinical Implications

Accurate classification of meningiomas using machine learning is an integral part of the clinic and is represented by systematic reviews of radiomics and MRI applications. Ugga et al. (2021) found that in their meta-analysis of 23 studies, machine learning (ML) techniques yielded an overall pooled AUC of 0.88 in predicting meningioma grade from preoperative MRIs, which demonstrates the potential of the application of imaging-based computational tools to assist clinical decisions. However, they also emphasized that although these radiomics models show great promise, they need to undergo copious amounts of validation and integration into standardized clinical workflows before they can be deployed in real-world neurosurgical decision-making. Methodological limitations and a lack of standardization currently prevent the translation of computational tools, such as the model in our study, from clinical applications.

MRI-based modeling has also been shown to aid in prognostic evaluation as well as grading. Hale et al. (2018) evaluated many ML algorithms on preoperative MRI features such as tumor volume, necrosis, edema and location, showing that neural networks gave an AUC of 0.8895 for the prediction of WHO grade II meningiomas. This predictive ability has direct implications for preoperative counseling as well as surgical planning because tumors of higher grade have a greater risk of recurrence and dictate the aggressiveness of resection strategies. Supporting this, Huntoon et al. (2020) reviewed the pathology of meningiomas and emphasized the heterogeneity of meningiomas stating that histological, molecular, and radiological features may vary widely regardless of being in the same WHO grade. This difference reinforces the clinical value of imaging-based ML system, which can better capture the subtle radiographic differences and patterns which may correspond to clinically significant subtypes.

Tumor localization has also been shown to be of further prognostic significance, affecting the feasibility of surgery and long-term outcome. Splavski et al. (2017) reviewed 243 patients with meningiomas and suggested a basic tumor localization scale, finding that peripheral tumor locations were linked with better outcomes than centrally located meningiomas in which neurovascular involvement makes resection more challenging. More recently, Hirschmann et al. (2025) found in a pediatric and adolescent cohort that tumor location is more determinative of relapse and outcome than WHO grade (skull base meningiomas had a 100% relapse rate as compared to 42% in other sites). Similarly, AlKhoshi et al (2023) demonstrated that postsurgical prognosis was highly dependent on tumor location especially if meningiomas occurred near critical skull base structures, while age and sex did not have a significant effect on prognosis. Together, these studies highlight how accurate preoperative classification and localization, which can be accomplished using transformer-based models, is not only academic but has direct implications for surgical planning, patient counseling and long-term prognosis.

### 5.3 Comparison with Conventional Deep Learning Approaches

Traditional ensemble and convolutional approaches have been known to be highly effective in brain tumor classification, yet they are significantly different from transformer-based architecture, especially due to their decreased robustness and adaptability. For example, M. et al. (2024) demonstrated that ensemble Deep Learning (DL) that integrated numerous CNN models instead of a single model reported notable improvement in sensitivity and specificity in MRI-based brain tumor classification. Although such approaches can mitigate overfitting and variance, their reliance on CNN-derived features limits their generalizability. This is especially true when compared to the self-attention mechanisms present in ViTs. Our fine-tuned ViT approach streamlines feature extraction using global dependencies across the MRI images, which adds robustness and resilience against variations in orientation and scale that ensemble CNNs typically are known to struggle with.

Single models such as CNNs, namely ResNet50, have been widely used as baselines for meningioma detection tasks. Singh and Prabha (2023) made a ResNet50-based classification pipeline using MRI data which achieved an accuracy of 96.63% with SoftMax activation and Adam optimization. Although this is an impressive accuracy, these architectures depend on convolutional kernels with limited receptive fields, which likely overlook long-range contextual information. On the other hand, our transformer pipeline models globally, improving performance when tumor margins are diffuse or heterogenous. These reasons highlight the increased effectiveness of ViTs when compared to conventional CNNs for more complex neuroimaging ML applications.

Finally, traditional ML models have also been explored extensively, providing a useful benchmark for evaluating the advantages of our deep learning. Al-Batah et al. (2025) compared six ML algorithms, including KNN, SVM, and Random Forests, across 15,000 MRI images of glioma, meningioma, and other tumors, finding that KNN and Neural Networks achieved classification accuracies of 98.5% and 98.4%, respectively. However, these methods would require intensive preprocessing, complex feature engineering, and do not generalize well beyond the training dataset. By contrast, transformer models automate feature learning, and they maintain robustness across varied data distributions. Reddy et al. (2024) reinforce this shift, showing that fine-tuned ViTs outperformed CNNs in multi-class brain tumor classification by capturing hierarchical patterns within MRI scans. Furthermore, Azam et al. (2025) introduced conical transformers to differentiate meningiomas from solitary fibrous tumors, achieving 94.68% accuracy in cross-validation and demonstrating strong robustness to rotation and scale variability in MRI scans. These findings show the architectural advantages of the transformer-based approach we employed in our pipeline.

### 5.4 Limitations of the Study

Our model displayed excellent classification of meningioma location however this alone is not enough to permit clinical use. Ugga et al. (2021) establishes that it is necessary to have large datasets for training deep learning models as models tend to not generalize as well with smaller datasets. Additionally, data consistency must be held to a high standard as low-quality data can lead to the lower accuracy in clinical use. However, even with larger, more consistent datasets there exists the issue of tumor heterogeneity which may challenge generalizations as seen by Huntoon et al. (2020). The dataset used in our study consisted of 1,094 meningioma MRI scans which may not be enough to be generalized to all populations clinically. A key challenge faced when training the model was the data scarcity which was approached by collating the original 29 classes into 11 classes as there were originally different classes for different weightings of the same anatomical region.

Besides limited data for training and evaluation, diverse data is necessary from multiple institutions using a myriad of MRI techniques. This concern must be addressed for true generalizability of a DL model to be used clinically in the future. Bento et al. (2022) highlights the growing need for the use large, multi-site, heterogenous brain imaging datasets for training, validation, and testing. Using such diverse datasets can yield for increased robustness of the model and lend to the ability to use such models in many hospitals despite different equipment or techniques. Variability in predictive accuracy for DL models when using data from different scanners is often referred to as domain shift. Mitigating domain shift is a particularly challenging problem for MRI imaging as it presents increased domain shift compared to X-ray or CT scans experimentally (Guo et al., 2024).

A key limitation of our study is the scope of our model only classifying location which does provide relevant clinical information but lacks the nuance required for treatment assessment, growth monitoring, and surgical planning. Boaro et al. (2022) explains that segmentation and volumetric analysis can provide such nuance and suffers less from inter-user variability compared the manual labeling. The study continues to demonstrate that a 3D-CNN could achieve a mean tumor segmentation Dice score of 85.2% and a median of 88.2%, comparable to inter-expert variability.

### 5.5 Future Directions and Improvements

Future work in meningioma classification should go beyond localization and additionally consider broader clinical outcomes. Musigmann et al. (2022) demonstrated that radiomics, paired with machine learning, can help predict surgical outcomes, such as the risk of subtotal resection, for example. By moving towards these broader clinical goals, future studies have the potential to not only identify tumor location, but to also anticipate treatment challenges. This would ultimately improve surgical planning and patient management, which would enhance the clinical relevance of AI classification models by combining diagnostic predictive capabilities with real-world decision making (Musigmann et al., 2022).

Improving on the success of the current Vision Transformers (ViTs) used, refinement of these transformer-based architectures is a promising next step. According to Reddy et al. (2024), fine-tuned ViTs significantly enhanced multi-class brain tumor classification performance, which demonstrates the potential improvements that may be made if architectural and training refinements tailored specifically to meningioma subtypes were made. Expanding datasets and adopting fine-tuning strategies optimized for tumor heterogeneity may help address limitations such as class imbalance (Reddy et al., 2024). This indicates that future iterations of meningioma classification models could greatly benefit from deeper and more specialized transformer designs than what were used in this instance.

Additionally, emerging transformer architectures should be investigated, as they hold promise for the neuro-oncological area of ML. Azam et al. (2025) introduced a conical transformer framework that was designed for rotation and scale invariance, and it achieved precise differentiation between meningiomas and solitary fibrous tumors, in this case. Innovations such as these could improve robustness against variability from MRI scans and extend the generalizability of meningioma classification models (Azan et al., 2025). Furthermore, combining ViTs with generative adversarial networks (GANs) could help overcome data scarcity and improve feature representation through GAN-based augmentation. Together, these directions highlight the need for architectural exploration and integrative modeling strategies that fuse classification, outcome prediction, and data expansion for stronger clinical translation (Huang et al., 2025).

The results highlight the effectiveness of Vision Transformers (ViTs) in meningioma location classification. The model’s ability to capture spatial dependencies enabled competitive performance with limited training data. However, class imbalance led to varying accuracy across tumor locations. This study underscores the importance of dataset diversity in medical AI applications. Future research should focus on expanding datasets and integrating ensemble learning strategies to improve classification reliability. Additionally, incorporating radiologist feedback for model validation may enhance clinical applicability.

## 6 Conclusion

Fine-Tuned Vision Transformers (ViTs) demonstrate strong potential for meningioma location classification from MRIs. By leveraging global attention mechanisms and spatial dependencies, the fine-tuned ViT achieved state-of-the-art level accuracy across multiple anatomical regions. Further work should address class imbalance and evaluate integration into radiologist workflows to assess clinical utility.

## Data Availability

All datasets and pre-trained machine learning models used in this study are open source. The code used for analysis in this study is not publicly archived but can be obtained upon request from the corresponding author.

https://huggingface.co/datasets/fernando2rad/neuro_cnn_meningioma_39c

## 7 Disclosures

The authors declare that there are no financial interests, commercial affiliations, or other potential conflicts of interest that could have influenced the objectivity of this research or the writing of this paper.

## Notes

### Competing Interest Statement

The authors have declared no competing interest.

### Funding Statement

This study did not receive any funding.

### Author Declarations

The source data, consisting of human MRI scan images of brains affected with meningioma, was publicly available before the initiation of the study and can be located at: https://huggingface.co/datasets/fernando2rad/neuro_cnn_meningioma_39c

